# Association of air pollution with brain health: A cross-sectional analysis in adults living in Canada

**DOI:** 10.1101/2025.10.21.25338504

**Authors:** Sandi M Azab, Sonia S Anand, Dany Doiron, Karleen M Schulze, Jeffrey R Brook, Michael Brauer, Dipika Desai, Matthias G Friedrich, Shrikant I Bangdiwala, Vikki Ho, Trevor J. B. Dummer, Paul Poirier, Jean-Claude Tardif, Koon K Teo, Scott Lear, Perry Hystad, Salim Yusuf, Eric E Smith, Russell J de Souza, the Canadian Alliance of Healthy Hearts and Minds (CAHHM) Study Investigators

## Abstract

**Background:** Air pollution is a risk factor for dementia but its role in early cognitive dysfunction is not clear. We aimed to investigate the association of air pollution with cognitive function, and the role of cardiovascular risk factors and greenspace in this association.

**Methods:** The Canadian Alliance for Healthy Hearts and Minds Cohort Study (CAHHM) is a cohort of Canadian adults recruited between 2014-2018, for whom averages of exposures to nitrogen dioxide (NO_2_) and fine particulate matter (PM_2.5_) were estimated for five years prior to recruitment. Outcomes included the *Montréal Cognitive Assessment* (MoCA) and *Digit Symbol Substitution Test (DSST)* for cognitive function, and magnetic resonance imaging-measured covert vascular brain injury. Generalized linear mixed models assessed pollutant associations with outcomes.

**Results:** A total of 6878 adults participated in the study with a mean age of 57.6 years (SD = 8.8) and 55.6% were women. Mean (SD; range) 5-year pollutant concentrations preceding enrolment (Figure S1) for PM_2.5_ was 6.9 μg/m³ (2.0; 1.8-11.2), and for NO_2_ was 12.9 ppb (5.9; 0.9-33.9). In adjusted models, a 5 μg/m^3^ higher PM_2.5_ concentration was associated with 0.44-points lower MoCA (95% confidence intervals (CI) −0.62, −0.25) and 1.31-points lower DSST (95% CI −2.41,-0.22) scores. A 5-ppb higher NO_2_ concentration was associated with 0.12-points lower MoCA (95% CI −0.17, −0.07) and 0.38 lower DSST (95% CI −0.70, −0.05) scores. A 5-ppb higher NO_2_ concentration was associated with higher odds of covert vascular brain injury (adjusted Odds Ratio (OR)=1.08; 95% CI 1.00, 1.17). Cardiovascular risk factors and greenspace did not change these associations.

**Conclusions:** PM_2.5_ and NO_2_ were associated with lower cognitive function scores in middle-aged adults living in Canada, independent of cardiovascular risk factors. This study suggests that air pollution mitigation efforts may help preserve cognitive function.

## 1. Introduction

Dementia is a major public health challenge (Hurd et al., 2013). There are currently 50 million people living with dementia worldwide and given increasing longevity, this number and associated healthcare costs are expected to rise (Delgado-Saborit et al., 2021). Along with lower educational attainment, smoking, hearing impairment, depression, infrequent social contact, traumatic brain injury, excessive alcohol consumption, hypertension, obesity, physical inactivity, and diabetes, air pollution is a modifiable risk factor for cognitive impairment and dementia (Livingston et al., 2020). However, the pathways underpinning this association are not well defined.

A recent systematic review and meta-analysis on ambient air pollution and clinical dementia (n=51 studies) suggested that exposure to fine particulate matter (PM_2.5_) was associated with increased risk of dementia (Wilker et al., 2023) with a pooled hazard ratio of 1.42 (95% confidence interval [CI]: 1.00 to 2.02 per 2 μg/m^3^ PM_2.5_) in a meta analysis of seven studies that used active case ascertainment, even at levels that met international air quality standards. A similar trend was found for nitrogen dioxide (NO_2_) although with more limited data (Wilker et al., 2023). Living close to heavy traffic (Chen et al., 2017b) and exposure to air pollutants (Chen et al., 2017a) were associated with a higher incidence of dementia in large population-based cohort studies in Canada (Smargiassi et al., 2020), a country with levels of air pollution among the lowest in the world (Weichenthal et al., 2022).

Oxidative stress, neuroinflammation, disruption of the blood-brain barrier (Block et al., 2004; Calderón-Garcidueñas et al., 2008), and worsening of cardiovascular risk factors (*e.g.,* obesity, hypertension, and diabetes) are potential mechanisms that underlie the association of air pollution with brain health. At the same time, it is proposed that greenspace can protect health via three main pathways of *reducing harm e.g.,* reduction in exposure to air pollution, *restoring capacity e.g.,* attention restoration, and *building capacity e.g.,* promoting social interactions (Markevych et al., 2017), yet, the evidence regarding the effect of greenspace on brain health is weak and conflicting (Delgado-Saborit et al., 2021; De Keijzer et al., 2016).

Dementia is preceded by decades of mostly subclinical cardiac and brain dysfunction (Power et al., 2016), therefore investigating the associations of chronic exposure to air pollution with cognitive function in middle-aged populations in a setting of low levels of air pollution e.g., Canada, may provide insight into early disease development. In middle-aged adults in The Canadian Alliance for Healthy Hearts and Minds Cohort (CAHHM) (Anand et al., 2016), we aimed to evaluate the association of exposure to PM_2.5_ and NO_2_ with cognitive function and covert vascular brain injury, and investigate the role of cardiovascular risk factors and neighbourhood greenspace in this association.

## 2. Methods

### 2.1. Study population

CAHHM is a prospective ‘cohort of cohorts’ study that includes 8580 adults from across Canada. Most participants (>80%) were recruited through existing cohorts between January 1, 2014 to December 31, 2018 from the provinces of British Columbia, Alberta, Ontario, Quebec, and Nova Scotia in mostly urban locations, as previously described (Anand et al., 2016). Ethics approval was obtained from the Hamilton Integrated Research Ethics Board (HiREB # 13-255), and all participants provided written informed consent. Participants were eligible if they were between ages 35 and 69 years at the time of enrollment into the parent cohort and willing to complete additional questionnaires, physical measurements, and a magnetic resonance imaging (MRI) scan. Participants were excluded if they had contraindications to an MRI scan, known history of clinical CVD (defined as a history of stroke, coronary artery disease, heart failure, or other heart disease), incomplete data on cardiovascular risk factors, or incomplete air pollutants values, resulting in a total of 6878 participants included in this study (**Figure 1**).

**Figure 1.**
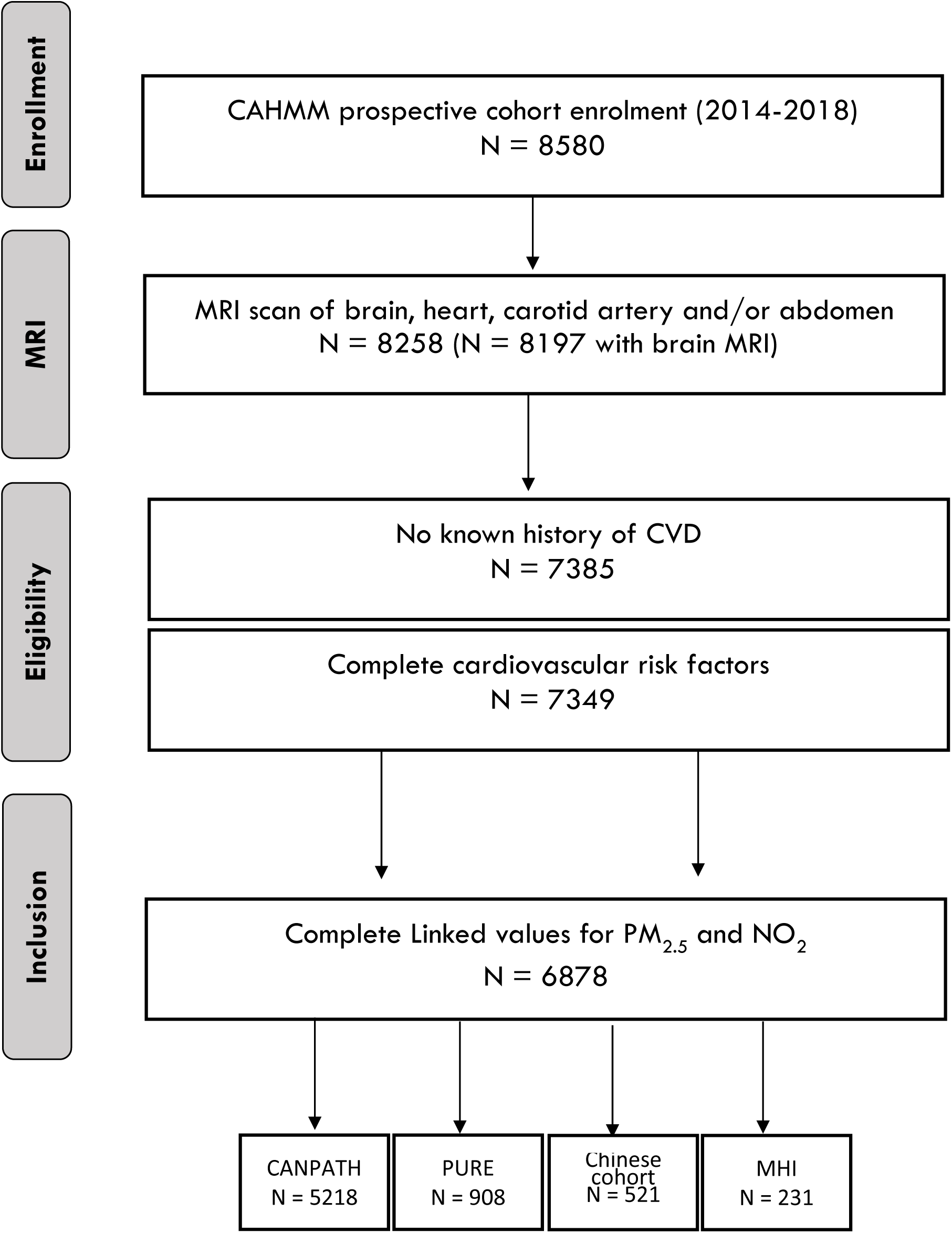
Flow chart for brain health measures and air pollution linkage in CAHHM

### 2.2. Measures of cognitive function

We used two measures of cognitive function. The Montreal Cognitive Assessment (MoCA) is a brief screening tool that evaluates delayed recall, verbal fluency, visuospatial skills, executive functions, calculation, abstraction, language, orientation, attention, and concentration (Nasreddine et al., 2005). Scores range from 0 to 30, and a score of 26 or higher denotes normal cognitive function. The Digital Symbol Substitution test (DSST) is a 2-minute test requiring participants to match symbols with numbers according to a code. Scores range from 0 to 133, and lower scores indicate worse performance (Kaufman, 1983). The DSST is commonly used in large international studies, because it is language-independent, sensitive, it can display age-related effects over the age of 40 to 70 years, and it can predict clinically important events including falls and mortality (Launer et al., 2011).

### 2.3. MRI-measured covert vascular brain injury

Details of the CAHHM MRI protocol have been previously published (Anand et al., 2016). The MRI scanners at each centre completed a validated test scan for quality assurance. Participants underwent a short non-contrast enhanced scan using a 1.5 Tesla or 3 Tesla magnet. To maximize MRI data comparability, a central core lab was used, procedures were defined in a manual of operations, sequence parameters were standardized and based on the validated Canadian Dementia Imaging Protocol, each MRI image was visually reviewed for quality, and visual reviews were performed by a limited group of qualified readers (two radiologists, with each scan read by one reader) who were required to meet core lab standards for inter-rater agreement (Duchesne et al., 2019). Brain infarcts were identified on a high resolution 3-dimensional T1-weighted sequence and 2-dimensional fluid-attenuated inversion recovery sequence by either magnetic resonance readers or a blinded neuroradiologist or neurologist. White matter hyperintensities (WMH) was rated on the Fazekas scale, a visual rating scale validated to correlate with volumetric measurements (Valdés Hernández et al., 2013). High WMH burden (HWMH) was defined as Fazekas score ≥4 (summing the periventricular and subcortical grades), which indicates beginning confluent or confluent WMH (Anand et al., 2016). Silent brain infarcts (SBIs) identified on MRI included small subcortical lacunes (<15 mm axial diameter), which are primarily related to small vessel disease, and larger (>15 mm axial diameter) or cortical infarcts, which may be caused by embolism (Anand et al., 2020). A covert vascular brain injury composite measure was analyzed, which included having either HWMH or one or more SBIs.

### 2.4. Assessment of ambient concentrations of PM_2.5_ and NO_2_

The primary air pollutant of interest was PM_2.5_ (fine inhalable particles ≤2.5 μm in aerodynamic diameter) and NO_2_ was a secondary measure; each representing different aspects of the air pollution mixture to which populations are exposed. The development of air pollution exposure datasets and distribution by the Canadian Urban Environmental Health Research Consortium (CANUE) (CANUE – The Canadian Urban Environmental Health Research Consortium et al., 2018) (www.canue.ca) have been documented previously (van Donkelaar et al., 2019; Hystad et al., 2011). Key emission sources for both primary and secondary PM_2.5_ are industrial emissions, wildfire smoke, space heating, residential wood heating, cooking, agriculture, and vehicle traffic emissions. Estimates of exposure to PM_2.5_ were derived across a 1×1 kilometer grid covering North America using NASA MODIS, MISR, and SeaWIFS satellite observations in combination with outputs from a global atmospheric chemical transport model (GEOS-Chem) (van Donkelaar et al., 2019). A geographically weighted regression (GWR) incorporating ground-based information on land cover, elevation, and aerosol composition adjusted for residual bias in the satellite-derived PM_2.5_ estimates (van Donkelaar et al., 2019). Comparison of PM_2.5_ estimates with in-situ monitors showed good agreement (R^2^ = 0.82) (Van Donkelaar et al., 2015).

NO_2_ is considered an indicator of urban traffic-related air pollution (TRAP), which is a complex mixture of gases and particles, including ultrafine particles (diameter ≤ 0.1 µm) proposed to reach the brain via the olfactory nerves (Cooper et al., 2020). Annual average NO_2_ concentrations in parts per billion (ppb) were estimated for each postal code location using a national land use regression (LUR) model for the year 2006 at 30 m resolution and adjusted for prior and subsequent years using long-term air quality monitoring station data (Hystad et al., 2011). The LUR NO_2_ model included road length, 2005-2011 satellite NO_2_ estimates, area of industrial land use within 2 km, and summer rainfall as predictors of regional NO_2_ variation (Hystad et al., 2011). Deterministic gradients were used to model local scale variation related to roads (i.e. traffic) (Hystad et al., 2011). The final NO_2_ model showed good performance, explaining 73% of the variation in measurements from national air pollution surveillance (NAPS) monitoring data with a root mean square error (RMSE) of 2.9 ppb.

Annual average PM_2.5_ and NO_2_ exposures for the five years prior to CAHHM recruitment (2008-2012), were linked to CAHHM using the six-character residential postal code of participants at the time of recruitment. We considered this exposure period representative of past long-term air pollution exposure concentrations prior to outcome assessment and relevant for investigating subclinical cognitive impairment, which manifests over a long period of time (Lusis, 2000).

### 2.5. Greenspace measure

We obtained residential greenspace estimates from CANUE measured as the mean of annual mean Normalised Difference Vegetation Index (NDVI) within 500 m buffer (representing greenspace within a 10-min walk) from residential postal codes. NDVI - a standard indicator of the intensity of green vegetation - uses difference in radiation between red and near-infrared wavelengths used by photosynthetic plants to absorb and emit radiation, respectively, to locate green vegetation (Srugo et al., 2019). Top of Atmosphere (TOA) reflectance data in bands from the United States Geological Survey’s Landsat 5 satellites were accessed via Google Earth Engine and CANUE staff used Google Earth Engine functions to create cloud free annual composites, and mask water features, then export the resulting band data to process into circular buffers (“Landsat 5 | U.S. Geological Survey,” n.d.; Gorelick et al., 2017) Greenspace was linked to CAHHM using the six-character residential postal code of participants at the time of recruitment to reflect the same exposure period as that of the pollutants. Values ranged from 0 (bare soil) to 1 (all green) and were categorized into tertiles.

### 2.6. Covariates

We conceptualized the set of potential confounders that were considered for multivariable models by carefully reviewing known risk factors for dementia, published associations and mechanisms, and by using diagrams. For education, participants were classified as having high school (or less), some post-secondary education (e.g., trade or technical training), and any college and/or a university earned certificate (bachelor’s or graduate degree). Employment status, income, and marital status were captured in the composite Social Disadvantage Index (SDI), a validated measure of individual-level socioeconomic status (low, moderate, high, and did not answer) (Anand et al., 2016) The following cardiovascular risk factors were collected using a validated tool(McGorrian et al., 2011; Yusuf et al., 2004): smoking status (never, former, and current smoker); physical activity (sedentary and leisure time active); depression (feeling sad, blue, or depressed for 2 weeks in a row in the past 12 months); home or work social stress (none, some, moderate, or high); central obesity (waist circumference per 10 cm); self-reported measures of history or treatment of diabetes (yes, no), hypertension (yes, no), and hyperlipidemia (yes, no).

### 2.7. Statistical analysis

The distribution of continuous variables is presented as means with standard deviation, and categorical variables are presented as counts and percentages. We assessed the association of the continuous measure of air pollutant exposure with the continuous outcomes of 1) MoCA and 2) DSST scores and with 3) the odds of having an MRI-detected covert vascular brain injury. Associations were calculated per 5 μg/m^3^ increment for PM_2.5_ and per 5 ppb increment for NO_2_, as relevant pollutant variation in high income countries (Wang et al., 2019; Boogaard et al., 2022). We used generalised linear mixed-effects models (GLMM) to explore associations and included a random intercept that represented the effect of recruitment centre. This random intercept served as a proxy for spatial clustering of participants across centre/neighbourhood and province. The following fixed covariates were included in each model, added progressively: 1) none (“unadjusted model”); 2) participant’s age, sex, and ethnicity (“basic model”); 3) established risk factors for dementia *i.e.,* education, smoking, and SDI (“risk factor model”); 4) lifestyle risk factors i.e., physical activity, depression and life stress (“lifestyle model”); 5) cardiovascular factors i.e. waist circumference, diabetes, hypertension, hyperlipidemia, and covert vascular brain injury when it was not the outcome (“maximal model”) and 6) greenspace (“greenspace model”). A complete case analysis was employed because missing data on outcomes and covariates were low (of n=6878: 0.06% for MoCA, 0.17% for DSST, 0.84% for covert vascular brain injury, 1.66% for education and 0.09% for greenspace). In sensitivity analyses, models 1-6 were 1) stratified by sex, 2) repeated after excluding recent immigrants to Canada of less than ten years (n=362), 3) repeated after excluding participants working outside their home community (n=2391) to address possible exposure assessment error due to major time away from residence, and 4) conducted in the subset of CAHHM participants with known history of clinical CVD (n=792) who were excluded from the study population.

An exploratory analysis was conducted to investigate the role of diabetes, hypertension, central obesity, and covert vascular brain injury in the association between air pollutants and cognitive scores in models adjusting for age, sex, ethnicity, and education (MacKinnon et al., 2007; Baron and Kenny, 1986) recognizing the inability to conduct a formal mediation analysis due to the lack of temporality between mediators and outcomes in the study design.

To investigate the role of greenspace, we added the greenspace variable as a covariate to the “maximal model 5”, as well as an interaction term between greenspace tertiles. If this interaction term was significant, it was considered evidence of interaction; and stratified models were presented.

A 2-sided p <0.05 was considered nominally significant with no adjustment for multiple testing. All analyses were completed using SAS software, version 9.4 (SAS Institute Inc).

### 2.8. Funding

No funder had a role in the data collection, analysis, interpretation, writing of the manuscript, or decision to submit.

## 3. Results

### 3.1. Participant characteristics

A total of 6878 adults participated in the study with a mean age of 57.6 years (SD = 8.8; range=32-81) and 55.6% were women (**Table 1**). 3372 (49.0%) participants were from Ontario, 1594 (23.2%) from Quebec, 774 (11.3%) from British Columbia, 720 (10.5%) from Nova Scotia, and 418 (6.1%) from Alberta. Over 97% of the cohort’s postal codes were in urban areas and mean (SD; range) 5-year pollutant concentrations preceding enrolment (Figure S1) for PM_2.5_ was 6.9 μg/m³ (2.0; 1.8-11.2), and for NO_2_ was 12.9 ppb (5.9; 0.9-33.9). The mean (SD) cognitive scores on the MoCA were 27.1 (2.3) and on the DSST were 74.2 (15.7)). On MRI scan 8.6% of study participants had covert vascular brain injury, of which 3.6% had silent brain infarcts, and 5.6% had HWMHs. Other demographic, anthropometric, and lifestyle characteristics of the study participants are found in **Table 1**.

**Table 1.** Characteristics of the study population.

|  | **N** | **Overall** |
| --- | --- | --- |
| Sex (female) | 6878 | 3823 (55.6) |
| Age, mean (SD), y | 6878 | 57.6 (8.8) |
| Self-reported ethnicity |  |  |
| East & South East Asian | 6878 | 944 (13.7) |
| South Asian | 6878 | 236 (3.4) |
| White Caucasian | 6878 | 5554 (80.8) |
| Other<sup>a</sup> | 6878 | 144 (2.1) |
| **Demographic, anthropometric, and lifestyle characteristics** |  |  |
| Highest Education Attained |  |  |
| High school or less | 6764 | 884 (13.1) |
| College or Trade | 6764 | 2171 (32.1) |
| University Degree | 6764 | 3709 (54.8) |
| Employment |  |  |
| Full or part time | 6760 | 4782 (70.7) |
| Retired | 6760 | 1480 (21.9) |
| No paid work | 6760 | 498 (7.4) |
| Household Income |  |  |
| <\$50,000 | 6353 | 1074 (16.9) |
| \$50-<\$75,000 | 6353 | 1165 (18.3) |
| \$75-<\$100,000 | 6353 | 1126 (17.7) |
| \$100-<\$150 000 | 6353 | 1538 (24.2) |
| \$150,000+ | 6353 | 1450 (22.8) |
| Income Not Reported | 6878 | 525 (7.6) |
| Living with partner/married | 6763 | 5114 (75.6) |
| Individual Social disadvantage score |  |  |
| Low disadvantage | 6334 | 3805 (60.1) |
| Moderate disadvantage | 6334 | 2142 (33.8) |
| High disadvantage | 6334 | 387 (6.1) |
| Smoking Status |  |  |
| Current (In past year) | 6878 | 364 (5.3) |
| Former (Quit >1 year ago) | 6878 | 2324 (33.8) |
| Never smoked | 6878 | 4190 (60.9) |
| Second Hand Smoke Exposure (1+ hrs/week) | 6878 | 319 (4.6) |
| Leisure Physical Inactivity | 6878 | 2705 (39.3) |
| Sad, blue or depressed for 2 weeks in past year | 6878 | 1158 (16.8) |

**Table 1. Characteristics of the study population**
|  | **N** | **Overall** |
| --- | --- | --- |
| INTERHEART risk score, mean (SD) | 6878 | 10.1 (5.8) |
| Low | 6878 | 3475 (50.5) |
| Moderate | 6878 | 2207 (32.1) |
| High | 6878 | 1196 (17.4) |
| Waist circumference, cm | 6878 | 88.7 (13.9) |
| Waist circumference obese, n (%) | 6878 | 2016 (29.3) |
| **Blood Pressure** |  |  |
| Systolic, mmHg | 6878 | 129 (17) |
| Diastolic, mmHg | 6878 | 80 (10) |
| **Medical History** |  |  |
| Diabetes on treatment | 6875 | 324 (4.7) |
| Elevated cholesterol &/or regular use of statins | 6878 | 2530 (36.8) |
| Regular use of statins | 6878 | 1184 (17.2) |
| Hypertension | 6876 | 2626 (38.2) |
| **Geographical characteristics** |  |  |
| Urban postal code | 6788 | 6615 (97.5) |
| Usual workplace location (Outside home community) | 6837 | 2391 (35.0) |
| **Provincial region, n (%)** |  |  |
| BC | 6878 | 774 (11.3) |
| AB | 6878 | 418 (6.1) |
| ON | 6878 | 3372 (49.0) |
| PQ | 6878 | 1594 (23.2) |
| NS | 6878 | 720 (10.5) |
| Home life or work stress in past year | 6878 | 2133 (31.0) |
| **Linked environmental measures** |  |  |
| PM<sub>2.5</sub>, ug/m<sup>3</sup>, over 2008-2012 | 6878 | 6.9 (2.0) |
| NO<sub>2</sub>, ppb, over 2008-2012 | 6878 | 12.9 (5.9) |
| Greenness Landsat Annual mean within 500m, over 2008-2011 | 6872 | 0.378 (0.091) |
| **Brain health characteristics** |  |  |
| MoCA (raw), mean (SD) | 6874 | 27.1 (2.3) |
| DSST, mean (SD) | 6866 | 74.2 (15.7) |
| Silent Brain Infarct, n (%) | 6828 | 245 (3.6) |
| Lacunar | 6828 | 158 (2.3) |

**Table 1. Characteristics of the study population**
|  | **N** | **Overall** |
| --- | --- | --- |
| Non-Lacunar (+/-lacunar) | 6828 | 87 (1.3) |
| High White Matter Hyperintensities, n (%) | 6839 | 384 (5.6) |
| Covert vascular Brain Injury<sup>b</sup>, n (%) | 6820 | 587 (8.6) |

Presented data are means (SD) unless otherwise indicated. <sup>a</sup>Includes Black people, Indigenous people, Mixed and unknown ethnicity people. <sup>b</sup>Covert vascular Brain Injury is a composite measure of silent brain infarcts and/or high white matter hyperintensities

### 3.2. Long-term pollutant exposure and cognitive outcomes

Associations of 5-year pollutant exposures with cognitive scores as measured by MoCA and DSST and with covert vascular brain injury as measured by MRI are presented in **Figure 2** and **Table 2**. Adjusting for potential confounders (Maximal model), a 5 μg/m^3^ increment in PM_2.5_ and a 5 ppb increment in NO_2_ concentrations were associated with 0.44-points lower (95% CI −0.62, −0.25; p<0.0001) and 0.12-points lower (95% CI −0.17, −0.07; p<0.0001) MoCA score, respectively. Similarly, a 5 μg/m^3^ increase in PM_2.5_ and a 5 ppb increase in NO_2_ concentrations were associated with 1.31-points lower (95% CI −2.41, −0.22; p=0.02) and 0.38-points lower (95% CI −0.70, −0.05; p=0.02) DSST score, respectively. While PM_2.5_ was not significantly associated with covert vascular brain injury (Odds Ratio (OR)=1.13 per 5 μg/m^3^; 95% confidence intervals (CI) 0.90, 1.42; p=0.24), each 5 ppb increment in NO_2_ was associated with 8% higher odds of covert vascular brain injury (OR=1.08; 95% CI 1.00, 1.17; p=0.04). The association of air pollutants with cognitive function persisted after adjustment for covert vascular brain injury, and the maximally adjusted models remained overall consistent in the sensitivity analysis among males and females (more so for MoCA than for DSST and covert vascular brain injury), after excluding immigrants with less than 10 years of residence in Canada, and after excluding participants working outside their home community (**Tables S1-S5**).

**Figure 2:**
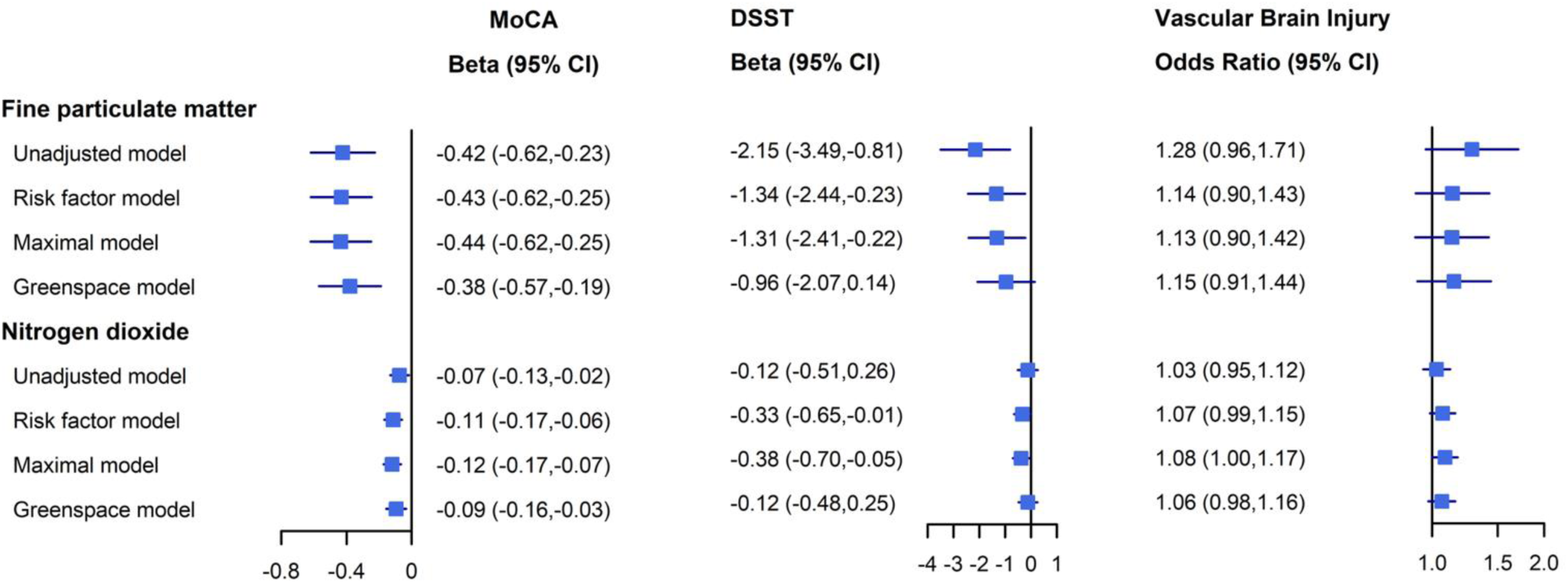
Associations of 5-year pollutant exposures with cognitive scores as measured by MoCA and DSST and with covert vascular brain injury (which included having either high white matter hyperintensities or one or more silent brain infarcts) as measured by MRI

**Table 2.** Associations of 5-year pollutant exposures with cognitive scores as measured by MoCA and DSST and with covert vascular brain injury as measured by MRI.

|  | MoCA |  | DSST |  | Covert vascular brain injury |  |
| --- | --- | --- | --- | --- | --- | --- |
| Model | Effect (95% CI) | P-value | Effect (95% CI) | P-value | Odds Ratio (95% CI) | P-value |
| **PM<sub>2.5</sub> per 5 µg/m<sup>3</sup>** |  |  |  |  |  |  |
| Model 1: No fixed effects | -0.42 (-0.62,-0.23) | <.0001 | -2.15 (-3.49,-0.81) | 0.002 | 1.28 (0.96,1.71) | 0.09 |
| Model 2: Age, sex, ethnicity | -0.41 (-0.60,-0.21) | <.0001 | -1.49 (-2.62,-0.36) | 0.01 | 1.13 (0.90,1.41) | 0.3 |
| Model 3: + Education, smoking, SDI | -0.44 (-0.62,-0.25) | <.0001 | -1.39 (-2.49,-0.28) | 0.01 | 1.13 (0.90,1.42) | 0.29 |
| Model 4: + physical activity, depression, life stress | -0.43 (-0.62,-0.25) | <.0001 | -1.34 (-2.44,-0.23) | 0.02 | 1.14 (0.90,1.43) | 0.27 |
| **Model 5: + cardiovascular factors** | **-0.44 (-0.62,-0.25)** | **<.0001** | **-1.31 (-2.41,-0.22)** | **0.02** | **1.13 (0.90,1.42)** | **0.29** |
| Model 6: + greenspace | -0.38 (-0.57,-0.19) | <.0001 | -0.96 (-2.07,0.14) | 0.09 | 1.15 (0.91,1.44) | 0.24 |
| **NO<sub>2</sub> per 5 ppb** |  |  |  |  |  |  |
| Model 1: No fixed effects | -0.07 (-0.13,-0.02) | 0.009 | -0.12 (-0.51,0.26) | 0.54 | 1.03 (0.95,1.12) | 0.5 |
| Model 2: Age, sex, ethnicity | -0.06 (-0.12,-0.01) | 0.03 | -0.20 (-0.53,0.13) | 0.24 | 1.07 (0.99,1.15) | 0.1 |
| Model 3: + Education, smoking, SDI | -0.11 (-0.17,-0.06) | <.0001 | -0.32 (-0.65,-0.00) | 0.05 | 1.07 (0.99,1.15) | 0.1 |
| Model 4: + physical activity, depression, life stress | -0.11 (-0.17,-0.06) | <.0001 | -0.33 (-0.65,-0.01) | 0.05 | 1.07 (0.99,1.15) | 0.1 |
| **Model 5: + cardiovascular factors** | **-0.12 (-0.17,-0.07)** | **<.0001** | **-0.38 (-0.70,-0.05)** | **0.02** | **1.08 (1.00,1.17)** | **0.04** |
| Model 6: + greenspace | -0.09 (-0.16,-0.03) | 0.002 | -0.12 (-0.48,0.25) | 0.53 | 1.06 (0.98,1.16) | 0.15 |

The following fixed covariates were included in each model progressively: 1) none (“unadjusted model”); 2) participant’s age, sex, and ethnicity (“basic model”); 3) further adjusted for established risk factors *i.e.*, education, smoking, and SDI (“risk factor model”); 4) further adjusted for lifestyle risk factors *i.e.*, physical activity, depression and life stress (“lifestyle model”); 5) further adjusted for cardiovascular factors *i.e.* waist circumference, diabetes, hypertension, hyperlipidemia, and covert vascular brain injury when it was not the outcome (“maximal model”) and 6) further adjusted for greenspace (“greenspace model”). SDI: social disadvantage index.

### 3.3. Role of cardiovascular risk factors

Results of single confounder/variable models to test whether central obesity, diabetes, hypertension, or MRI-detected vascular brain injury could be transmitting the effects of PM_2.5_ and NO_2_ on lower cognitive scores are presented in **Figure 3**. Firstly, a significant relation of the exposures to the outcomes was established. Next, we observed a significant relation of the exposures to the four hypothesized mediating variables only in the models for NO_2_ with hypertension and central obesity (waist circumference). Hypertension and central obesity were significantly related to both MoCA and DSST when added to a model with NO_2_. Lastly, there was no change in the NO_2_ effect on MoCA or DSST when adjusting for hypertension or central obesity. Thus, based on these empirical models, we conclude that neither diabetes, hypertension, central obesity, or covert vascular brain injury changed the effect of air pollution on cognitive function.

**Figure 3:**
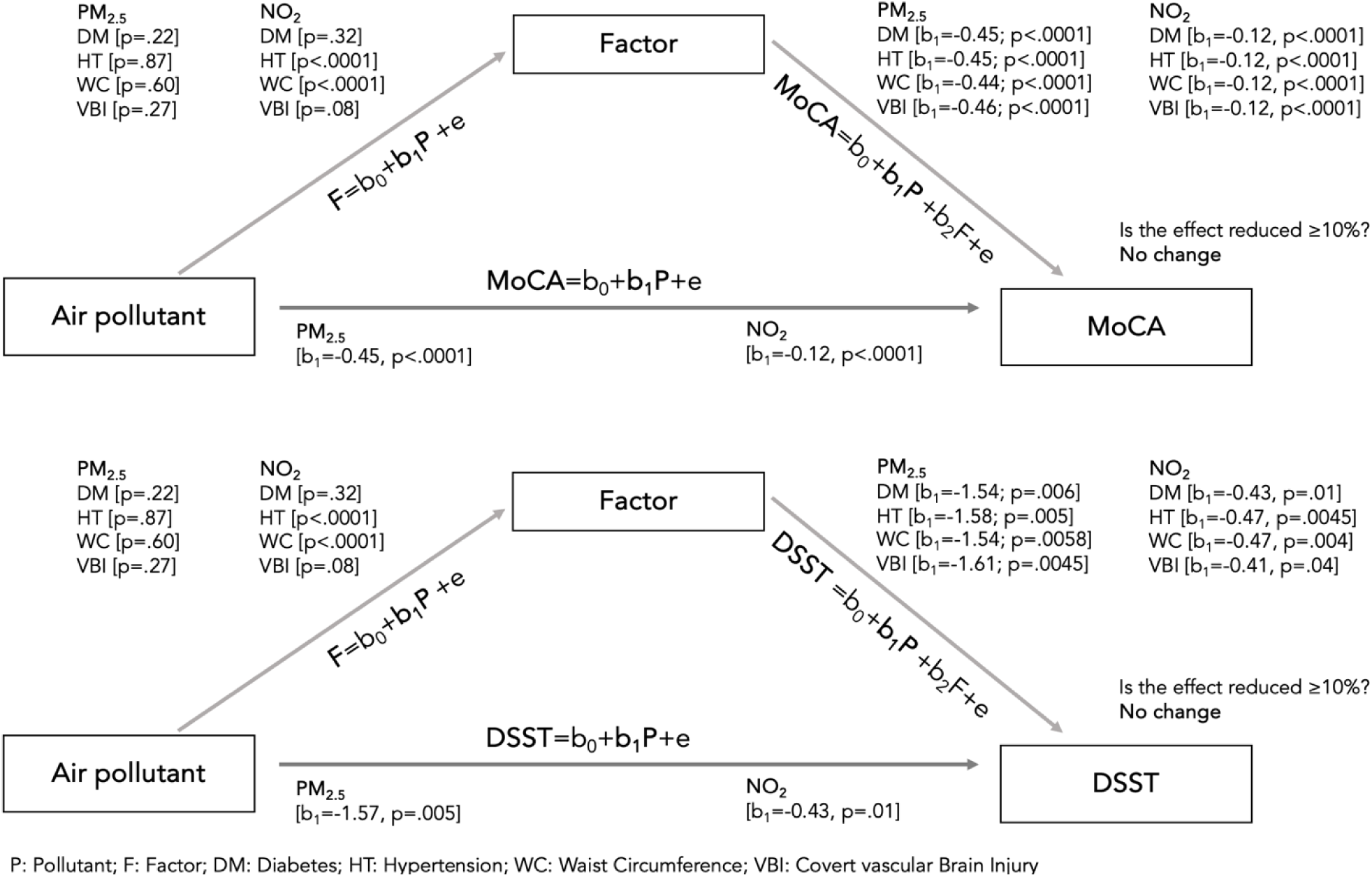
Exploratory analysis of pollutant exposures and cardiovascular risk factors with cognitive scores as measured by MoCA and DSST

### 3.4. Greenspace

Adding greenspace to the models (model 6) did not materially change the estimates (**Figure 2** and **Table 2**). When adding an interaction term to test whether the effect of the air pollutants on cognitive function changes when greenspace exposure changes, p-value from F-tests was not significant except for NO_2_ and MoCA (p-interaction <0.02) and NO_2_ and DSST (p-interaction < 0.02). From stratified analysis we concluded that greenspace did not blunt the effect of air pollution on cognitive function (**Table S6**),

## 4. Discussion

In this cohort of 6,878 middle-aged adults living in Canada, where levels of air pollution are amongst the lowest in the world, long-term exposure to ambient air pollution was consistently associated with reduced cognitive function as measured by MoCA and DSST. These findings were consistent among men and women for MoCA and remained robust after exclusion of recent immigrants and those working outside of their residential neighbourhood. Further investigation of these findings in populations who may be most vulnerable e.g., children and the elderly is needed. We examined various covariates and confounders, and found that diabetes, hypertension, central obesity, and covert vascular brain injury as measured by MRI, did not change the effect of air pollution on cognitive function despite their known impact on cognition.

In 2021, the World Health Organization updated its air quality guidelines lowering the recommended annual levels of PM_2.5_ from ten to five μg/m³ and NO_2_ from 21 to five ppb (K. Zhang et al., 2023). In 2020, the Canadian Council of Ministers of the Environment updated the Canadian Ambient Air Quality Standards (CAAQS) recommended updated annual levels of PM_2.5_ from ten (2015) to 8.8 μg/m³ and NO_2_ to 17 ppb (Brauer et al., 2022). In the current study, both air pollutants, PM_2.5_ [mean (SD) = 6.9 μg/m³ (2.0); IQR=3.54] and NO_2_ [mean (SD) = 12.9 ppb (5.9); IQR=9.68] were associated with worse performance on both MoCA, a test of global cognition across multiple domains such as memory and verbal comprehension, and DSST, a measure of processing speed that demands visual scanning, attention, and working memory. A 5 μg/m^3^ increase in PM_2.5_ and a 5 ppb increase in NO_2_ concentrations associated estimates could be interpreted as equivalent to 12.6 and 3.4 years of cognitive aging, respectively, for MoCA, and as equivalent to 1.9 and 0.5 years of cognitive aging, respectively, for DSST. These equivalents were calculated by extrapolating the estimates of the pollutants compared to the estimates of age (by 10-year increments) in the maximal models. Prior publications point to associations between air pollution and lower global cognition, as well as impairment of specific cognitive domains (Delgado-Saborit et al., 2021); more conclusively for PM_2.5_ than for other pollutants (Weuve et al., 2021). For instance, the National Social Life, Health, and Aging Project (NSHAP) reported a PM_2.5_ 1-year inter-quartile range-increase (4.25 μg/m^3^) was associated with reduced global cognition equivalent to 1.6 years of aging in older adults across the United States (Tallon et al., 2017). [mean (SD) =13.07 μg/m³ (7.23)]. A prior analysis of the UK Biobank cohort suggested that PM_2.5_ was associated with slower reaction time although follow-up analysis did not show an association with changes in cognitive score [median (q1, q3) = 9.86 (9.32, 10.40)] (Cullen et al., 2018). Longitudinal studies of air pollution and cognitive decline showed either significant (Kilian and Kitazawa, 2018), weak (Cullen et al., 2018), or no effects [median (q1, q3) = 13.6 (12.2, 14.8)] (Loop et al., 2013) and evidence remains inconclusive (Slomski, 2022). However, living in locations in the US with improvement in air quality over 10 years was associated with slower decline in cognitive function in older women (equivalent to 0.9-1.6 years younger) (Younan et al., 2022) highlighting potential benefits of action aimed at reducing ambient air pollution (Slomski, 2022).

There is a major gap in the knowledge of the causal pathway from air pollution exposure to cognitive decline and dementia. Wilker et al. used the ROBINS-E tool specifically focused on environmental exposures to assess risk of bias in studies of air pollution and dementia and the confounding domain was a reported threat (Wilker et al., 2023). Identifying all confounders in the relationship between air pollution and cognitive outcomes is difficult as much remains still unknown (Delgado-Saborit et al., 2021). In our multivariable models, we have attempted to investigate individual-level factors comprehensively including some that have limited prior study, such as depression, stress, physical activity, and greenspace, and those factors that are proposed mediators (Delgado-Saborit et al., 2021). The absence of a strong dampening of association when central obesity, diabetes, hypertension, or covert vascular brain injury on MRI were adjusted for in CAHHM, is further supported by a US cohort study of 27,857 individuals which found that hypertension and stroke were not mediating or modifying the association of PM_2.5_ with dementia (B. Zhang et al., 2023). This suggests presence of some direct effect not mediated by cardiovascular factors or vascular brain injury, tentatively through systemic inflammation and biochemical perturbations, neuroinflammation (toxic microglia activation), and increased oxidative stress (Calderón-Garcidueñas et al., 2008; Block and Calderón-Garcidueñas, 2009, Costa et al., 2017). Additional hypotheses include sex differences and gene-environment interactions. Exposure to ultrafine particles (<0.1 μm), which are difficult to measure or regulate, may also contribute to oxidative stress and inflammation in the CNS, which could start very early in life (Andrea Flood-Garibay et al., 2023).

CAHHM is one of the largest studies of heart and brain health worldwide, with individual-level deep phenotyping and MRI of the brain, heart, carotid, and abdomen in persons aged 35-69 linked to postal code-level pollutant concentrations, which enabled us to study the early relationship of long-term exposure to air pollution with subclinical cognitive dysfunction that typically precedes for decades prior to a dementia diagnosis (Azab et al., 2024; Anand et al., 2016). We have previously shown in CAHHM that no postsecondary education, moderate and high levels of cardiovascular risk factors, and MRI vascular brain injury accounted for 15%, 19%, and 10% of low cognitive test scores, respectively (Anand et al., 2020). We have also found that visceral adipose tissue accounted for 20% of reduced DSST score, independently, in this cohort (Anand et al., 2022). Although effect estimates of air pollution are smaller than other risk factors, policy changes can be powerful given the ubiquitous extent of populations’ passive exposure to air pollutants (Livingston et al., 2020; Wilker et al., 2023). As long as there is no cure for dementia, acting on acknowledged modifiable risk factors remains the most feasible strategy to reduce incidence and progression.

Our study has the following limitations. Individual-level exposures were estimated based on baseline residence address; thus, exposure misspecification is possible because our study could not consider commuting, indoor, or occupational exposures. Nevertheless, residential address remains a key exposure site for populations (Brasche and Bischof, 2005). Because of the lack of residential history and increasingly less precise air pollution exposure data further back in time, we assumed that the restricted timeframe of exposure at baseline residential address is representative of past exposures, and we were not able to conduct time-varying analyses. Future investigations are needed to examine varying exposure time-windows and lag periods. Next, the 5-year pollutant exposure period was fixed for all participants (2008-2012) regardless of when an individual’s enrolment occurred in the 2014-2018 window. However, studies have demonstrated temporal stability in the spatial patterns of air pollutants over 10 years (Wang et al., 2013). Despite our extensive attempt to control for confounders, some covariates were unmeasured such as traffic noise, hearing impairment, excessive alcohol consumption, and head injury, and the risk of residual confounding cannot be excluded. Lastly, due to the observational nature of the study, causality cannot be established and reverse causation, though very unlikely, cannot be ruled out.

## 5. Conclusion

Our results indicate a consistent association between air pollution and cognitive function in middle-aged adults in Canada, independent of typical cardiovascular risk factors. This warrants further studies on the impact of strategies to reduce exposure to air pollution on cognitive function, quality of life, and outcomes.

## Data Availability

CAHHM data cannot be deposited publicly as these collaborative data originate from multiple Canadian cohorts with different legal frameworks. CAHHM represents a "cohort of cohorts", and at the time participants were enrolled into each respective cohort, data sharing was not part of the consent process. Thus, the appropriate legal frameworks to allow for data to be deposited publicly (identified or de-identified) is not in place, nor was such sharing anticipated or run by the respective REB at the time of cohort invitations. Therefore, any requests for data may be made to CAHHM, with requests for data sharing to be considered on a case-by-case basis. A detailed statistical analysis plan and the code used in the analysis are also available on reasonable request.

## Acknowledgements

**Steering Committee of Canadian Alliance of Healthy Hearts and Minds:** S.S. Anand (Chair)*, M.G. Friedrich (Co-Chair), Douglas S. Lee (Co-Chair), P Awadalla (Ontario Health Study), T. Dummer (BC Generations Project), J. Vena (Alberta’s Tomorrow Project), P. Broët (CARTaGENE), J. Hicks (Atlantic PATH), J-C. Tardif (MHI Biobank), K. Teo, S. Yusuf (PURE-Central), B-M. Knoppers (Ethics, Legal and Social Issues). **Project Office Staff at Population Health Research Institute (PHRI):** D. Desai, S. Zafar. **Statistics/Biometrics Programmers Team at PHRI:** K. Schulze, S. Bangdiwala. **Cohort Operations Research Personnel:** K McDonald (Ontario Health Study), N. Noisel (CARTaGENE), J. Chu (BC Generations Project), J. Hicks (Atlantic PATH), H. Whelan (Alberta’s Tomorrow Project), S. Rangarajan (PURE), D. Busseuil (MHI Biobank). **Site Investigators and Staff:** J. Leipsic, S. Lear, V. de Jong; M. Noseworthy, K. Teo, E. Ramezani, N. Konyer; P. Poirier, A-S. Bourlaud, E Larose, K. Bibeau; J. Leipsic, S. Lear, V. de Jong; E. Smith, R. Frayne, A. Charlton, R Sekhon; A. Moody, V. Thayalasuthan; A.Kripalani, G Leung; M. Noseworthy, S. Anand, R. de Souza, N. Konyer, S. Zafar; G. Paraga,L. Reid; A.J. Dick, F. Ahmad; D. Kelton, H. Shah; F. Marcotte, H. Poiffaut; M.G. Friedrich, J. Lebel; E. Larose, K. Bibeau; R. Miller, L. Parker, D. Thompson, J. Hicks; J-C. Tardif, H.Poiffaut; J. Tu, K. Chan, A. Moody, V. Thayalasuthan. **MRI Working Group and Core Lab Investigators/Staff:** Chair: M.G. Friedrich; Brain Core Lab: E.E. Smith; Carotid Core Lab: A. Moody, V. Thayalasuthan; Abdomen: E. Larose, K. Bibeau, Cardiac: F. Marcotte, F. Henriques. **Contextual Working Group:** R. de Souza, S. Anand, G. Booth, J. Brook, D. Corsi, L. Gauvin, S. Lear, F. Razak, S.V. Subramanian, J. Tu. **CAHHM Founding Advisory Group:** Jean Rouleau, Pierre Boyle, Caroline Wong, Eldon Smith.

The Canadian Alliance of Healthy Hearts and Minds (CAHHM) investigators include the following individuals: Sonia S. Anand, MD, PhD, Department of Medicine, Department of Health Research Methods, Evidence, and Impact, McMaster University, Population Health Research Institute, Hamilton Health Sciences, Hamilton, Canada; Philip Awadalla, PhD, Department of Molecular Genetics, Ontario Institute for Cancer Research, University of Toronto, Toronto, Canada; Sandra E. Black, MA, MD, OC, Department of Medicine (Neurology), Sunnybrook Health Sciences Centre, University of Toronto, Toronto, Canada; Broët Philippe, MD, PhD, Department of Preventive and Social Medicine, École de santé publique, Université de Montréal, and Research Centre, CHU Sainte Justine, Montréal, Canada; Alexander Dick, MD, Department of Medicine, University of Ottawa Heart Institute, Ottawa, Canada; Trevor Dummer, PhD, MSc, Department of Epidemiology, Biostatistics, and Public HealthPractice, School of Population and Public Health, University of British Columbia, and BC Cancer Agency, Vancouver, Canada; Matthias G. Friedrich, MD, Department of Cardiology, McGill University, Montréal, Canada; Jason Hicks, MSc, Atlantic Partnership for Tomorrow’s Health, Dalhousie University, Halifax, Canada; David Kelton, MD, RPVI, Department of Medicine, William Osler Health System; Anish Kirpalani, MD, MASc, Department of Medical Imaging, St. Michael’s Hospital, University of Toronto, Toronto, Canada; Maria BarthaKnoppers, PhD, Centre of Genomics and Policy, McGill University, Montréal, Canada; Scott A. Lear, PhD, Department of Pathology, Simon Fraser University, Burnaby, Canada; Eric Larose, DVM, MD, Department of Medicine, University of Laval, Quebec City, Canada; Russell J. de Souza, RD, ScD, Department of Health Research Methods, Evidence, and Impact, McMaster University, Hamilton, Canada; Douglas S. Lee, MD, PhD, ICES Central, Cardiovascular Research Program, Institute for Clinical Evaluative Sciences, Peter Munk Cardiac Centre University Health Network, Department of Medicine, University of Toronto, Toronto, Canada; Jonathan Leipsic, MD, Department of Radiology, University of British Columbia, Vancouver, Canada; Francois Marcotte, MD, Department of Cardiology, Montreal Heart Institute, University of Montreal, Montréal, Canada; Alan R. Moody, MBBS, Department of Medical Imaging, University of Toronto, Sunnybrook Health Sciences Centre, Toronto, Canada; Michael D. Noseworthy, PhD, PEng, Department of Electrical and Computer Engineering, McMaster University, St. Joseph’s Health Care, Hamilton, Canada; Grace Parraga, PhD, Department of Medical Biophysics, Robarts Research Institute, Western University, London, Canada; Louise Parker, PhD, Atlantic Partnership for Tomorrow’s Health, Dalhousie University, Halifax, Canada; Paul Poirier, MD, PhD, Institut de cardiologie et de pneumologie de Quebec, Université of Laval, Quebec City, Canada; Eric E. Smith, MD, MPH, Hotchkiss Brain Institute, Department of Clinical Neurosciences, University of Calgary, Calgary, Canada; Jean-Claude Tardif, MD, Department of Cardiology, Montreal Heart Institute, University of Montreal, Montréal, Canada; Koon K. Teo, MBBCh, PhD, Department of Medicine, Department of Health Research Methods, Evidence, and Impact, McMaster University, Population Health Research Institute, Hamilton Health Sciences, Hamilton, Canada; Jack V. Tu, MD, PhD, MSc, Department of Medicine, University of Toronto, Institute for Clinical Evaluative Sciences, Sunnybrook Schulich Heart Centre, Toronto, Canada (deceased); Jennifer Vena, PhD, Cancer Research and Analytics, Cancer Control Alberta, Alberta Health Services, Edmonton, Canada.

The Canadian Partnership for Tomorrow’s Health (CanPath) research is only possible with the commitment of its research participants, its staff and its funders. The data and/or biosamples used in this research were made available by CanPath and the British Columbia Generations Project, Alberta’s Tomorrow Project, Ontario Health Study, CARTaGENE, and Atlantic Partnership for Tomorrow’s Health.

## CRediT authorship contribution statement

**Sandi M Azab:** Writing – original draft, Writing – review & editing, Conceptualization, Formal analysis, Visualization. **Sonia S Anand:** Writing – review & editing, Funding acquisition, Conceptualization, Investigation, Supervision. **Dany Doiron:** Writing – review & editing, Methodology, Conceptualization. **Karleen M Schulze:** Formal analysis, Data curation, Visualization, Methodology, Writing – review & editing. **Jeffrey R Brook:** Writing – review & editing, Data curation, Funding acquisition, Conceptualization, Methodology, Investigation. **Michael Brauer:** Writing – review & editing, Conceptualization, Investigation, Supervision. **Dipika Desai:** Writing – review & editing, Project administration, Data curation. **Matthias G Friedrich:** Writing – review & editing, Funding acquisition, Investigation. **Shrikant I Bangdiwala:** Writing – review & editing, Methodology, Formal analysis, Investigation. **Vikki Ho:** Writing – review & editing, Investigation. **Trevor J. B. Dummer:** Writing – review & editing, Investigation. **Paul Poirier:** Writing – review & editing, Investigation. **Jean-Claude Tardif:** Writing – review & editing, Investigation. **Koon K Teo:** Writing – review & editing, Investigation. **Scott Lear:** Writing – review & editing, Investigation. **Perry Hystad:** Writing – review & editing, Investigation **Salim Yusuf:** Writing – review & editing, Funding acquisition, Investigation. **Eric E Smith:** Writing – review & editing, Funding acquisition, Conceptualization, Investigation, Supervision. **Russell J de Souza:** Writing – review & editing, Methodology, Funding acquisition, Conceptualization, Investigation, Supervision.

---

**Figure S1.**
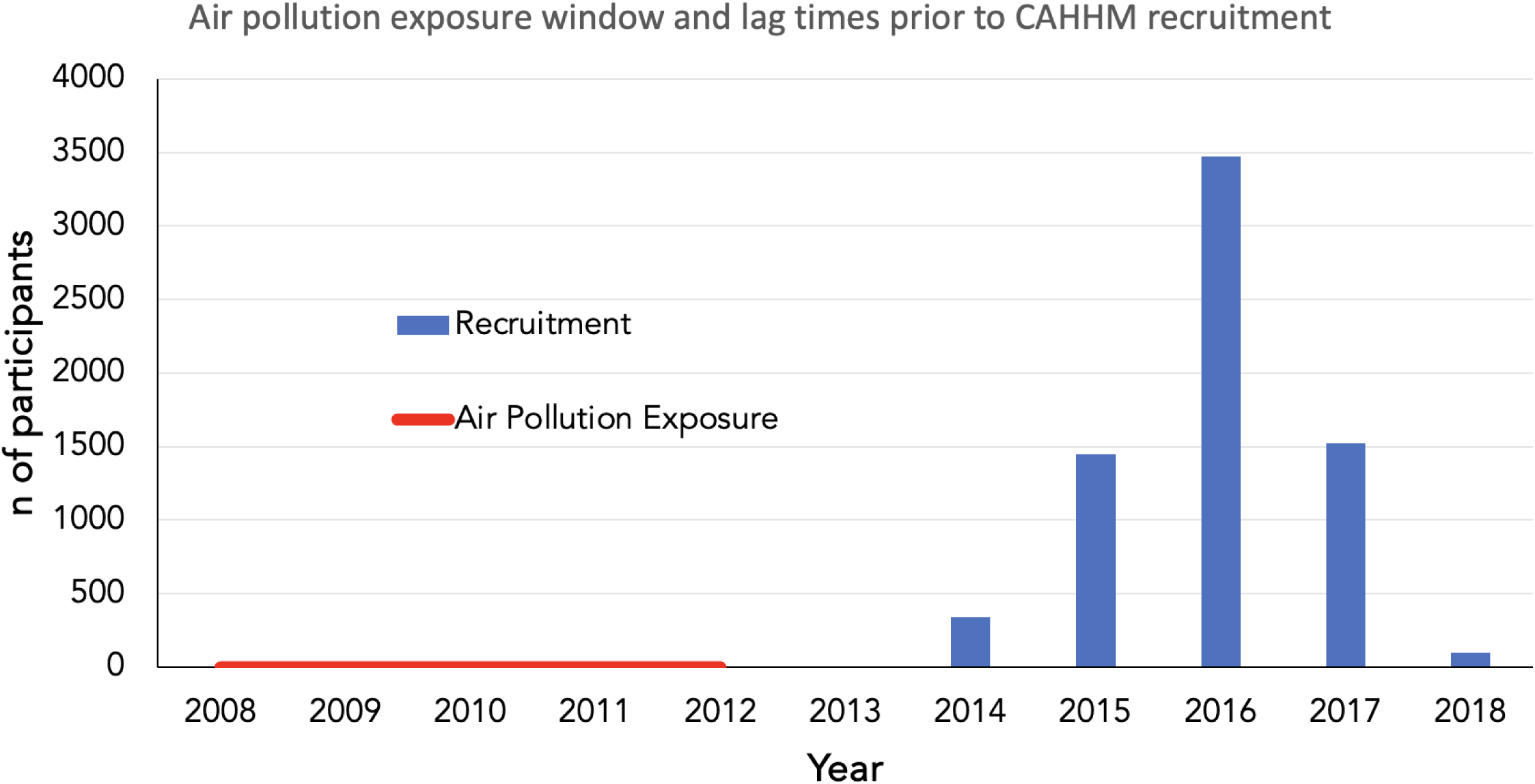
Temporal outline of the exposure measurement (5-year average of chronic air pollution exposure) preceding the start of recruitment and outcome assessment in CAHHM cohort with lag time ranging from 1 to 5 years.

**Table S1.** Sensitivity analysis of the associations of 5-year pollutant exposures with MoCA.

|  | Females (N=3722) |  | Males (N=2975) |  | Recent immigrants excluded (N=6338) |  | Working outside of community excluded (N=4353) |  |
| --- | --- | --- | --- | --- | --- | --- | --- | --- |
| Model | Effect (95% CI) | P-value | Effect (95% CI) | P-value | Effect (95% CI) | P-value | Effect (95% CI) | P-value |
| **PM<sub>2.5</sub> per 5 µg/m<sup>3</sup>** |  |  |  |  |  |  |  |  |
| Model 1: No fixed effects | -0.36 (-0.60,-0.11) | 0.005 | -0.47 (-0.77,-0.17) | 0.002 | -0.46 (-0.66,-0.26) | <.0001 | -0.37 (-0.61,-0.12) | 0.003 |
| Model 2: Age, sex, ethnicity | -0.34 (-0.59,-0.09) | 0.008 | -0.42 (-0.72,-0.13) | 0.005 | -0.41 (-0.61,-0.21) | <.0001 | -0.39 (-0.63,-0.15) | 0.001 |
| Model 3: + Education, smoking, SDI | -0.37 (-0.62,-0.13) | 0.003 | -0.45 (-0.74,-0.16) | 0.002 | -0.44 (-0.63,-0.25) | <.0001 | -0.41 (-0.64,-0.18) | <0.001 |
| Model 4: + physical activity, depression, life stress | -0.37 (-0.61,-0.13) | 0.003 | -0.45 (-0.74,-0.17) | 0.002 | -0.43 (-0.63,-0.24) | <.0001 | -0.41 (-0.65,-0.18) | <0.001 |
| **Model 5: + cardiovascular factors** | **-0.37 (-0.61,-0.13)** | **0.003** | **-0.48 (-0.77,-0.20)** | **<0.001** | **-0.43 (-0.63,-0.24)** | **<.0001** | **-0.41 (-0.64,-0.17)** | **<0.001** |
| Model 6: + greenspace | -0.32 (-0.57,-0.08) | 0.01 | -0.41 (-0.70,-0.12) | 0.006 | -0.38 (-0.58,-0.18) | <0.001 | -0.35 (-0.59,-0.12) | 0.004 |
| **NO<sub>2</sub> per 5 ppb** |  |  |  |  |  |  |  |  |
| Model 1: No fixed effects | -0.07 (-0.14,0.00) | 0.06 | -0.06 (-0.15,0.03) | 0.18 | -0.07 (-0.13,-0.01) | 0.02 | -0.10 (-0.17,-0.03) | 0.005 |
| Model 2: Age, sex, ethnicity | -0.05 (-0.12,0.02) | 0.16 | -0.06 (-0.15,0.02) | 0.16 | -0.05 (-0.11,0.00) | 0.06 | -0.09 (-0.16,-0.02) | 0.01 |
| Model 3: + Education, smoking, SDI | -0.12 (-0.18,-0.05) | 0.001 | -0.10 (-0.18,-0.01) | 0.03 | -0.11 (-0.16,-0.05) | <0.001 | -0.14 (-0.21,-0.08) | <.0001 |
| Model 4: + physical activity, depression, life stress | -0.12 (-0.19,-0.05) | <0.001 | -0.10 (-0.18,-0.02) | 0.02 | -0.11 (-0.16,-0.05) | <0.001 | -0.14 (-0.21,-0.08) | <.0001 |
| **Model 5: + cardiovascular factors** | **-0.12 (-0.19,-0.05)** | **<0.001** | **-0.11 (-0.19,-0.02)** | **0.01** | **-0.11 (-0.17,-0.06)** | **<.0001** | **-0.15 (-0.22,-0.08)** | **<.0001** |
| Model 6: + greenspace | -0.12 (-0.20,-0.04) | 0.003 | -0.05 (-0.15,0.04) | 0.27 | -0.09 (-0.15,-0.02) | 0.007 | -0.14 (-0.22,-0.06) | <0.001 |

The following fixed covariates were included in each model progressively: 1) none ("unadjusted model"); 2) participant's age, sex, and ethnicity ("basic model"); 3) further adjusted for established risk factors *i.e.*, education, smoking, and SDI ("risk factor model"); 4) further adjusted for lifestyle risk factors *i.e.*, physical activity, depression and life stress ("lifestyle model"); 5) further adjusted for cardiovascular factors *i.e.* waist circumference, diabetes, hypertension, and hyperlipidemia and covert silent vascular brain injury ("maximal model") and 6) further adjusted for greenspace ("greenspace model"). SDI: social disadvantage index.

**Table S2.** Sensitivity analysis of the associations of 5-year pollutant exposures with DSST.

|  | Females (N=3720) |  | Males (N=2970) |  | Recent immigrants excluded (N=6331) |  | Working outside of community excluded (N=4347) |  |
| --- | --- | --- | --- | --- | --- | --- | --- | --- |
| Model | Effect (95% CI) | P-value | Effect (95% CI) | P-value | Effect (95% CI) | P-value | Effect (95% CI) | P-value |
| **PM<sub>2.5</sub> per 5 µg/m<sup>3</sup>** |  |  |  |  |  |  |  |  |
| Model 1: No fixed effects | -3.11 (-4.84,-1.38) | <0.001 | -0.91 (-2.83,1.02) | 0.36 | -2.64 (-4.03,-1.25) | <0.001 | -1.79 (-3.44,-0.14) | 0.03 |
| Model 2: Age, sex, ethnicity | -2.17 (-3.65,-0.68) | 0.004 | -0.36 (-2.00,1.27) | 0.66 | -1.67 (-2.84,-0.51) | 0.005 | -1.63 (-3.01,-0.25) | 0.02 |
| Model 3: + Education, smoking, SDI | -2.05 (-3.52,-0.58) | 0.006 | -0.26 (-1.83,1.31) | 0.75 | -1.57 (-2.71,-0.42) | 0.007 | -1.49 (-2.84,-0.14) | 0.03 |
| Model 4: + physical activity, depression, life stress | -1.97 (-3.43,-0.50) | 0.008 | -0.22 (-1.78,1.34) | 0.79 | -1.52 (-2.66,-0.38) | 0.009 | -1.41 (-2.76,-0.06) | 0.04 |
| **Model 5: + cardiovascular factors** | **-1.91 (-3.35,-0.46)** | **0.01** | **-0.30 (-1.85,1.25)** | **0.7** | **-1.47 (-2.60,-0.34)** | **0.01** | **-1.35 (-2.69,-0.02)** | **0.05** |
| Model 6: + greenspace | -1.64 (-3.10,-0.18) | 0.03 | -0.02 (-1.55,1.52) | 0.98 | -1.10 (-2.24,0.04) | 0.06 | -1.06 (-2.40,0.27) | 0.12 |
| **NO<sub>2</sub> per 5 ppb** |  |  |  |  |  |  |  |  |
| Model 1: No fixed effects | -0.38 (-0.87,0.11) | 0.12 | 0.44 (-0.12,1.01) | 0.13 | -0.15 (-0.55,0.24) | 0.45 | -0.07 (-0.55,0.40) | 0.76 |
| Model 2: Age, sex, ethnicity | -0.37 (-0.80,0.06) | 0.09 | 0.13 (-0.37,0.63) | 0.6 | -0.18 (-0.51,0.15) | 0.29 | -0.19 (-0.59,0.22) | 0.36 |
| Model 3: + Education, smoking, SDI | -0.48 (-0.91,-0.06) | 0.03 | 0.02 (-0.46,0.51) | 0.92 | -0.32 (-0.66,0.01) | 0.05 | -0.34 (-0.75,0.06) | 0.1 |
| Model 4: + physical activity, depression, life stress | -0.49 (-0.91,-0.06) | 0.03 | 0.01 (-0.47,0.49) | 0.97 | -0.33 (-0.66,0.00) | 0.05 | -0.32 (-0.72,0.08) | 0.11 |
| **Model 5: + cardiovascular factors** | **-0.57 (-0.99,-0.15)** | **0.008** | **-0.00 (-0.48,0.48)** | **0.99** | **-0.37 (-0.70,-0.04)** | **0.03** | **-0.37 (-0.77,0.03)** | **0.07** |
| Model 6: + greenspace | -0.41 (-0.89,0.07) | 0.09 | 0.36 (-0.17,0.88) | 0.19 | -0.11 (-0.48,0.27) | 0.58 | -0.19 (-0.63,0.25) | 0.4 |

The following fixed covariates were included in each model progressively: 1) none ("unadjusted model"); 2) participant's age, sex, and ethnicity ("basic model"); 3) further adjusted for established risk factors *i.e.*, education, smoking, and SDI ("risk factor model"); 4) further adjusted for lifestyle risk factors *i.e.*, physical activity, depression and life stress ("lifestyle model"); 5) further adjusted for cardiovascular factors *i.e.* waist circumference, diabetes, hypertension, and hyperlipidemia and covert silent vascular brain injury ("maximal model") and 6) further adjusted for greenspace ("greenspace model"). SDI: social disadvantage index.

**Table S3.** Sensitivity analysis of the associations of 5-year pollutant exposures with covert vascular brain injury.

|  | Females (N=3724) |  | Males (N=2977) |  | Recent immigrants excluded (N=6342) |  | Working outside of community excluded (N=4357) |  |
| --- | --- | --- | --- | --- | --- | --- | --- | --- |
| Model | Odds Ratio (95% CI) | P-value | Odds Ratio (95% CI) | P-value | Odds Ratio (95% CI) | P-value | Odds Ratio (95% CI) | P-value |
| **PM<sub>2.5</sub> per 5 µg/m<sup>3</sup>** |  |  |  |  |  |  |  |  |
| Model 1: No fixed effects | 1.52 (1.12,2.05) | 0.007 | 0.94 (0.66,1.35) | 0.74 | 1.25 (0.94,1.65) | 0.12 | 1.14 (0.83,1.55) | 0.42 |
| Model 2: Age, sex, ethnicity | 1.43 (1.05,1.96) | 0.02 | 0.86 (0.62,1.20) | 0.38 | 1.12 (0.89,1.41) | 0.33 | 1.16 (0.90,1.50) | 0.26 |
| Model 3: + Education, smoking, SDI | 1.48 (1.08,2.02) | 0.02 | 0.85 (0.61,1.19) | 0.34 | 1.12 (0.89,1.41) | 0.32 | 1.18 (0.91,1.53) | 0.21 |
| Model 4: + physical activity, depression, life stress | 1.51 (1.10,2.08) | 0.01 | 0.84 (0.61,1.17) | 0.31 | 1.13 (0.90,1.42) | 0.31 | 1.18 (0.91,1.53) | 0.22 |
| **Model 5: + cardiovascular factors** | **1.51 (1.10,2.08)** | **0.01** | **0.83 (0.59,1.16)** | **0.27** | **1.12 (0.89,1.41)** | **0.33** | **1.17 (0.90,1.51)** | **0.24** |
| Model 6: + greenspace | 1.52 (1.10,2.08) | 0.01 | 0.85 (0.61,1.19) | 0.36 | 1.14 (0.90,1.43) | 0.27 | 1.18 (0.91,1.53) | 0.21 |
| **NO<sub>2</sub> per 5 ppb** |  |  |  |  |  |  |  |  |
| Model 1: No fixed effects | 1.00 (0.91,1.11) | 0.95 | 1.05 (0.93,1.18) | 0.43 | 1.03 (0.95,1.12) | 0.41 | 1.05 (0.96,1.16) | 0.28 |
| Model 2: Age, sex, ethnicity | 1.03 (0.93,1.14) | 0.61 | 1.13 (1.01,1.26) | 0.04 | 1.07 (0.99,1.15) | 0.1 | 1.09 (1.00,1.19) | 0.05 |
| Model 3: + Education, smoking, SDI | 1.03 (0.93,1.15) | 0.58 | 1.13 (1.01,1.27) | 0.04 | 1.06 (0.98,1.15) | 0.12 | 1.09 (1.00,1.19) | 0.05 |
| Model 4: + physical activity, depression, life stress | 1.03 (0.93,1.15) | 0.57 | 1.13 (1.01,1.27) | 0.03 | 1.07 (0.99,1.15) | 0.11 | 1.09 (1.00,1.19) | 0.05 |
| **Model 5: + cardiovascular factors** | **1.04 (0.94,1.16)** | **0.44** | **1.16 (1.03,1.30)** | **0.02** | **1.08 (1.00,1.17)** | **0.05** | **1.11 (1.02,1.21)** | **0.02** |
| Model 6: + greenspace | 1.03 (0.92,1.17) | 0.58 | 1.12 (0.99,1.26) | 0.08 | -0.11 (-0.48,0.27) | 0.58 | 1.10 (1.00,1.20) | 0.05 |

The following fixed covariates were included in each model progressively: 1) none ("unadjusted model"); 2) participant's age, sex, and ethnicity ("basic model"); 3) further adjusted for established risk factors *i.e.*, education, smoking, and SDI ("risk factor model"); 4) further adjusted for lifestyle risk factors *i.e.*, physical activity, depression and life stress ("lifestyle model"); 5) further adjusted for cardiovascular factors *i.e.* waist circumference, diabetes, hypertension, and hyperlipidemia ("maximal model") and 6) further adjusted for greenspace ("greenspace model"). SDI: social disadvantage index.

**Table S4.**
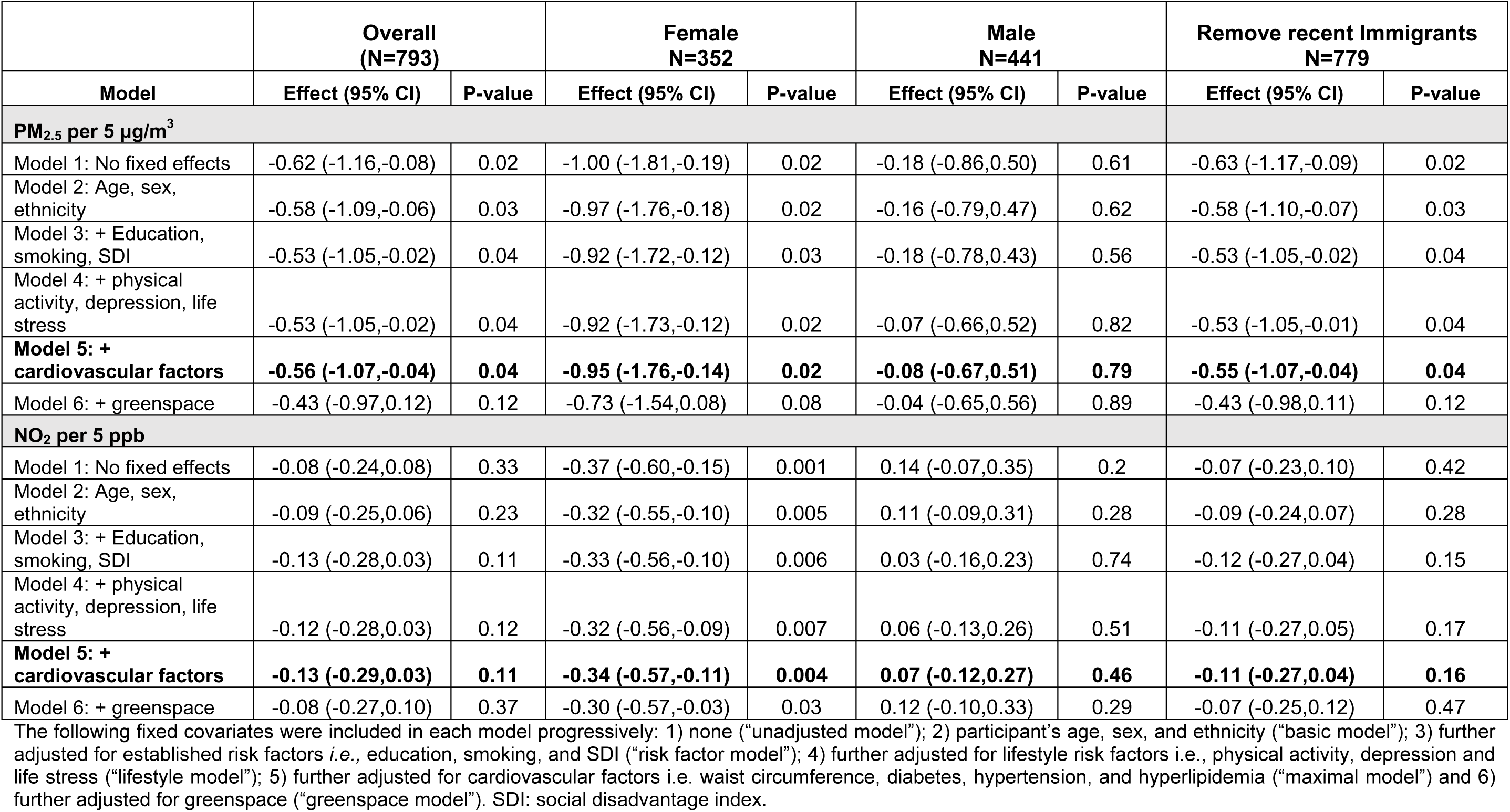
Sensitivity analysis of the associations of 5-year pollutant exposures with MoCA in the subset with clinical CVD.

|  | **Overall**<br>**(N=793)** |  | **Female**<br>**N=352** |  | **Male**<br>**N=441** |  | **Remove recent Immigrants**<br>**N=779** |  |
| --- | --- | --- | --- | --- | --- | --- | --- | --- |
| **Model** | **Effect (95% CI)** | **P-value** | **Effect (95% CI)** | **P-value** | **Effect (95% CI)** | **P-value** | **Effect (95% CI)** | **P-value** |
| **PM<sub>2.5</sub> per 5 µg/m<sup>3</sup>** |  |  |  |  |  |  |  |  |
| Model 1: No fixed effects | -0.62 (-1.16,-0.08) | 0.02 | -1.00 (-1.81,-0.19) | 0.02 | -0.18 (-0.86,0.50) | 0.61 | -0.63 (-1.17,-0.09) | 0.02 |
| Model 2: Age, sex, ethnicity | -0.58 (-1.09,-0.06) | 0.03 | -0.97 (-1.76,-0.18) | 0.02 | -0.16 (-0.79,0.47) | 0.62 | -0.58 (-1.10,-0.07) | 0.03 |
| Model 3: + Education, smoking, SDI | -0.53 (-1.05,-0.02) | 0.04 | -0.92 (-1.72,-0.12) | 0.03 | -0.18 (-0.78,0.43) | 0.56 | -0.53 (-1.05,-0.02) | 0.04 |
| Model 4: + physical activity, depression, life stress | -0.53 (-1.05,-0.02) | 0.04 | -0.92 (-1.73,-0.12) | 0.02 | -0.07 (-0.66,0.52) | 0.82 | -0.53 (-1.05,-0.01) | 0.04 |
| **Model 5: + cardiovascular factors** | **-0.56 (-1.07,-0.04)** | **0.04** | **-0.95 (-1.76,-0.14)** | **0.02** | **-0.08 (-0.67,0.51)** | **0.79** | **-0.55 (-1.07,-0.04)** | **0.04** |
| Model 6: + greenspace | -0.43 (-0.97,0.12) | 0.12 | -0.73 (-1.54,0.08) | 0.08 | -0.04 (-0.65,0.56) | 0.89 | -0.43 (-0.98,0.11) | 0.12 |
| **NO<sub>2</sub> per 5 ppb** |  |  |  |  |  |  |  |  |
| Model 1: No fixed effects | -0.08 (-0.24,0.08) | 0.33 | -0.37 (-0.60,-0.15) | 0.001 | 0.14 (-0.07,0.35) | 0.2 | -0.07 (-0.23,0.10) | 0.42 |
| Model 2: Age, sex, ethnicity | -0.09 (-0.25,0.06) | 0.23 | -0.32 (-0.55,-0.10) | 0.005 | 0.11 (-0.09,0.31) | 0.28 | -0.09 (-0.24,0.07) | 0.28 |
| Model 3: + Education, smoking, SDI | -0.13 (-0.28,0.03) | 0.11 | -0.33 (-0.56,-0.10) | 0.006 | 0.03 (-0.16,0.23) | 0.74 | -0.12 (-0.27,0.04) | 0.15 |
| Model 4: + physical activity, depression, life stress | -0.12 (-0.28,0.03) | 0.12 | -0.32 (-0.56,-0.09) | 0.007 | 0.06 (-0.13,0.26) | 0.51 | -0.11 (-0.27,0.05) | 0.17 |
| **Model 5: + cardiovascular factors** | **-0.13 (-0.29,0.03)** | **0.11** | **-0.34 (-0.57,-0.11)** | **0.004** | **0.07 (-0.12,0.27)** | **0.46** | **-0.11 (-0.27,0.04)** | **0.16** |
| Model 6: + greenspace | -0.08 (-0.27,0.10) | 0.37 | -0.30 (-0.57,-0.03) | 0.03 | 0.12 (-0.10,0.33) | 0.29 | -0.07 (-0.25,0.12) | 0.47 |

The following fixed covariates were included in each model progressively: 1) none ("unadjusted model"); 2) participant's age, sex, and ethnicity ("basic model"); 3) further adjusted for established risk factors *i.e.*, education, smoking, and SDI ("risk factor model"); 4) further adjusted for lifestyle risk factors *i.e.*, physical activity, depression and life stress ("lifestyle model"); 5) further adjusted for cardiovascular factors *i.e.* waist circumference, diabetes, hypertension, and hyperlipidemia ("maximal model") and 6) further adjusted for greenspace ("greenspace model"). SDI: social disadvantage index.

**Table S5.** Sensitivity analysis of the associations of 5-year pollutant exposures with DSST in the subset with clinical CVD.

|  | Overall<br>N=792 |  | Female<br>N=351 |  | Male<br>N=441 |  | Remove recent Immigrants<br>N=778 |  |
| --- | --- | --- | --- | --- | --- | --- | --- | --- |
| Model | Effect (95% CI) | P-value | Effect (95% CI) | P-value | Effect (95% CI) | P-value | Effect (95% CI) | P-value |
| **PM<sub>2.5</sub> per 5 µg/m<sup>3</sup>** |  |  |  |  |  |  |  |  |
| Model 1: No fixed effects | -2.36 (-5.81,1.09) | 0.18 | -5.06 (-10.1,-0.07) | 0.05 | 0.88 (-3.37,5.13) | 0.68 | -2.37 (-5.80,1.07) | 0.18 |
| Model 2: Age, sex, ethnicity | -1.49 (-4.36,1.37) | 0.31 | -3.32 (-7.71,1.06) | 0.14 | 0.40 (-3.20,4.00) | 0.83 | -1.33 (-4.16,1.51) | 0.36 |
| Model 3: + Education, smoking, SDI | -0.67 (-3.39,2.04) | 0.63 | -2.12 (-6.33,2.10) | 0.32 | 0.82 (-2.53,4.17) | 0.63 | -0.53 (-3.23,2.17) | 0.7 |
| Model 4: + physical activity, depression, life stress | -0.76 (-3.44,1.92) | 0.58 | -2.20 (-6.38,1.98) | 0.3 | 0.85 (-2.45,4.15) | 0.61 | -0.64 (-3.31,2.03) | 0.64 |
| **Model 5: + cardiovascular factors** | **-0.97 (-3.64,1.70)** | **0.47** | **-2.36 (-6.51,1.80)** | **0.27** | **0.49 (-2.82,3.80)** | **0.77** | **-0.84 (-3.49,1.81)** | **0.53** |
| Model 6: + greenspace | -1.07 (-3.90,1.76) | 0.46 | -0.83 (-4.87,3.21) | 0.69 | -0.67 (-4.28,2.94) | 0.72 | -1.05 (-3.88,1.77) | 0.46 |
| **NO<sub>2</sub> per 5 ppb** |  |  |  |  |  |  |  |  |
| Model 1: No fixed effects | 0.34 (-0.67,1.36) | 0.51 | -0.56 (-1.98,0.87) | 0.44 | 0.89 (-0.42,2.19) | 0.18 | 0.38 (-0.64,1.40) | 0.47 |
| Model 2: Age, sex, ethnicity | -0.09 (-0.95,0.77) | 0.83 | -0.38 (-1.65,0.88) | 0.55 | 0.46 (-0.69,1.60) | 0.43 | -0.04 (-0.90,0.82) | 0.93 |
| Model 3: + Education, smoking, SDI | -0.17 (-1.00,0.66) | 0.69 | -0.21 (-1.45,1.02) | 0.73 | 0.14 (-0.95,1.24) | 0.8 | -0.12 (-0.94,0.71) | 0.78 |
| Model 4: + physical activity, depression, life stress | -0.13 (-0.95,0.69) | 0.76 | -0.24 (-1.47,0.99) | 0.7 | 0.17 (-0.91,1.26) | 0.75 | -0.07 (-0.89,0.75) | 0.87 |
| **Model 5: + cardiovascular factors** | **-0.24 (-1.06,0.58)** | **0.56** | **-0.38 (-1.60,0.84)** | **0.54** | **-0.01 (-1.09,1.08)** | **0.99** | **-0.16 (-0.98,0.66)** | **0.7** |
| Model 6: + greenspace | -0.34 (-1.30,0.62) | 0.49 | 0.57 (-0.72,1.86) | 0.39 | -0.63 (-1.90,0.64) | 0.33 | -0.30 (-1.26,0.67) | 0.54 |

The following fixed covariates were included in each model progressively: 1) none ("unadjusted model"); 2) participant's age, sex, and ethnicity ("basic model"); 3) further adjusted for established risk factors *i.e.*, education, smoking, and SDI ("risk factor model"); 4) further adjusted for lifestyle risk factors *i.e.*, physical activity, depression and life stress ("lifestyle model"); 5) further adjusted for cardiovascular factors *i.e.* waist circumference, diabetes, hypertension, and hyperlipidemia ("maximal model") and 6) further adjusted for greenspace ("greenspace model"). SDI: social disadvantage index.

**Table S6.** Stratified analysis of the associations of 5-year NO_2_ exposure (per 5 ppb) with MoCA and DSST within low, medium, and high greenspace tertiles.

|  | Overall |  | Low greenspace |  | Medium greenspace |  | High greenspace |  |
| --- | --- | --- | --- | --- | --- | --- | --- | --- |
| Model | Effect (95% CI) | P-value | Effect (95% CI) | P-value | Effect (95% CI) | P-value | Effect (95% CI) | P-value |
| **MoCA** |  |  |  |  |  |  |  |  |
| Model 1: No fixed effects | -0.07 (-0.13,-0.02) | 0.0095 | 0.15 (0.03,0.27) | 0.0115 | -0.11 (-0.22,0.01) | 0.0632 | -0.14 (-0.24,-0.03) | 0.0131 |
| Model 2: Age, sex, ethnicity | -0.06 (-0.12,-0.01) | 0.0292 | 0.14 (0.03,0.26) | 0.0128 | -0.09 (-0.20,0.02) | 0.1208 | -0.09 (-0.19,0.02) | 0.1118 |
| Model 3: + Education, smoking, SDI | -0.11 (-0.17,-0.06) | <.0001 | 0.09 (-0.02,0.20) | 0.1132 | -0.15 (-0.26,-0.04) | 0.0074 | -0.17 (-0.27,-0.07) | 0.0010 |
| Model 4: + physical activity, depression, life stress | -0.11 (-0.17,-0.06) | <.0001 | 0.08 (-0.03,0.19) | 0.1495 | -0.15 (-0.26,-0.04) | 0.0086 | -0.17 (-0.28,-0.07) | 0.0010 |
| **Model 5: + cardiovascular factors** | **-0.12 (-0.17,-0.07)** | **<.0001** | **0.07 (-0.03,0.18)** | **0.1789** | **-0.17 (-0.28,-0.05)** | **0.0036** | **-0.17 (-0.28,-0.07)** | **0.0008** |
| **DSST** |  |  |  |  |  |  |  |  |
| Model 1: No fixed effects | -0.12 (-0.51,0.26) | 0.5359 | 0.05 (-0.76,0.87) | 0.8977 | 0.30 (-0.49,1.08) | 0.4582 | -0.02 (-0.76,0.72) | 0.9622 |
| Model 2: Age, sex, ethnicity | -0.20 (-0.53,0.13) | 0.2373 | -0.13 (-0.80,0.55) | 0.7142 | 0.29 (-0.34,0.93) | 0.3693 | 0.44 (-0.18,1.07) | 0.1642 |
| Model 3: + Education, smoking, SDI | -0.32 (-0.65,-0.00) | 0.0497 | -0.21 (-0.86,0.45) | 0.5389 | 0.11 (-0.51,0.72) | 0.7338 | 0.09 (-0.52,0.70) | 0.7803 |
| Model 4: + physical activity, depression, life stress | -0.33 (-0.65,-0.01) | 0.0461 | -0.20 (-0.86,0.45) | 0.5404 | 0.10 (-0.51,0.72) | 0.7421 | 0.08 (-0.52,0.69) | 0.7864 |
| **Model 5: + cardiovascular factors** | **-0.38 (-0.70,-0.05)** | **0.0220** | **-0.27 (-0.92,0.37)** | **0.4095** | **0.01 (-0.61,0.62)** | **0.9848** | **0.01 (-0.60,0.61)** | **0.9829** |

The following fixed covariates were included in each model progressively: 1) none ("unadjusted model"); 2) participant's age, sex, and ethnicity ("basic model"); 3) further adjusted for established risk factors *i.e.*, education, smoking, and SDI ("risk factor model"); 4) further adjusted for lifestyle risk factors *i.e.*, physical activity, depression and life stress ("lifestyle model"); 5) further adjusted for cardiovascular factors *i.e.* waist circumference, diabetes, hypertension, hyperlipidemia and covert vascular brain injury ("maximal model"). SDI: social disadvantage index.

